# Risk and protective factors against cognitive decline in older adults from a nationally representative sample in India: Results from the LASI-DAD

**DOI:** 10.1101/2025.07.07.25331027

**Authors:** Alden L. Gross, Emma Nichols, Miguel Arce Rentería, Pranali Y. Khobragade, Erik Meijer, Mary Ganguli, Eileen M. Crimmins, Sharmistha Dey, Aparajit Ballav Dey, Jinkook Lee, LASI-DAD Authorship Team

## Abstract

**INTRODUCTION:** We characterized modifiable risk and protective factors for cognitive decline in India.

**METHODS:** Using the first nationally representative population-based longitudinal sample of N=6,168 older adults in India, we evaluated associations of risk factors (demographic characteristics, self-reported and objective health characteristics, health behaviors, and sensory function) for late-life cognitive decline with up to 6.4 years of follow-up (range: 2.8 to 6.4 years).

**RESULTS:** The mean rate of general cognitive decline was -0.029 SD per year, and was progressively steeper with age. Most risk factors, particularly demographic and cardiovascular characteristics, were associated with steeper cognitive decline in expected directions: associations of history of high cholesterol or heart attack on rate of cognitive decline, for example, were comparable to being 11-12 years older.

**DISCUSSION:** Most risk factors were associated with change in expected directions, highlighting the potential generalizability to India of previously identified risk factors for dementia.

## Introduction

Progressive, irreversible cognitive decline, whether due to either Alzheimer Disease or other pathologies, is a hallmark of dementia.^(1)^ Longitudinal assessments of cognition are thus essential to the study of dementia, and prior research has emphasized the importance of longitudinal cognitive data to establish robust associations of risk and protective factors for dementia.^(2–4)^ Studying cognitive decline over time allows us to better inform prognosis by determining which factors exacerbate the rate of cognitive decline, particularly since absolute level of cognitive performance may be explained by earlier life experiences such as education, occupation, and work experience, and a plethora of related sequelae through the lifecourse.^(5)^ Although promising cross-sectional associations with cognitive outcomes may suggest potential causal relationships by providing a snapshot of potential associations,^(6)^ such associations must be replicated using data on cognitive decline to establish temporal precedence of risk and protective factors.^(8)^ Longitudinal data are especially important given the long prodromal periods of most dementias. Despite the recognized importance of cognitive change, for logistical and other reasons, longitudinal data on cognitive performance among large, nationally representative samples are not widely available outside high-income countries. To address this gap and to enable longitudinal assessments of cognition and dementia in India, the second wave of the Harmonized Diagnostic Assessment of Dementia for the Longitudinal Aging Study in India (LASI-DAD) was fielded.^(7)^

Despite its value to uncovering causes of dementia, longitudinal cognitive data from lower- and middle-income countries, including India, are comparatively rare. With over 1.4 billion people today, India is the most populous country in the world, and with rapid population aging, the number of people aged 60 years and older is projected to reach 347 million by 2050, accounting for over 20% of the world population aged 60 and older.^(9.10)^ Longitudinal assessments of comprehensive cognitive performance using population-representative data in India represent an important innovation, particularly given that current knowledge outside of high-income countries is limited and moreover that prior longitudinal studies in India have been based on geographically restricted or otherwise non-representative clinical or convenience samples.

The goal of the present study was to evaluate the sociodemographic and health predictors of cognitive decline in a nationally representative sample in India. To evaluate correlates of cognitive change, we considered a variety of modifiable and non-modifiable risk factors previously shown to be associated with cognitive outcomes, spanning demographic factors, health history, health and function, health behaviors, and sensory function. We used late-life cognitive decline at the second wave to study a series of modifiable and non-modifiable risk factors measured prior to the first wave. To address measurement changes in the LASI-DAD cognitive battery, we leveraged confirmatory factor analysis models to cocalibrate general and domain-specific cognitive factors across waves.

## Methods

### Study sample

We used data from the first two waves of LASI-DAD. LASI-DAD began as a substudy of the Longitudinal Aging Study in India (LASI), which recruited a nationally representative sample of N=73,408 adults aged 45+ years from 29 states and 6 union territories in India from 2017-18 using a stratified, multistage area probability sample.^(11)^ Wave 1 of LASI-DAD recruited N=4,096 participants in 2017-20 from LASI using a stratified random sampling strategy, such that the sample is nationally representative of India.^(12)^ In wave 2 of LASI-DAD, conducted 2022-24, N=2,566 participants from wave 1 and a refresher sample of N=2,072 were recruited using a similar sampling strategy, for a nationally representative total of N=4638 in wave 2. At the beginning of wave 1, participants in LASI-DAD were interviewed at a local hospital. Later in wave 1 and throughout wave 2, participants were interviewed in their home. Ethics approval was obtained from the Indian Council of Medical Research (2202-16741/F1) and all collaborating institutions. Consent materials were translated into 12 languages.

Participants provided written informed consent. For participants unable to read, consent forms were read aloud by interviewers, and participants unable to sign their name made a mark on a tablet screen witnessed by a legally authorized representative.

### Variables

Key variables used for this study were cognitive test items from LASI-DAD waves 1 and 2, as well as 30 risk factors and variables assessed in the core LASI survey commonly found to be associated with cognition. These included measures of demographic characteristics, health and everyday function, measures of objective physical activity, health behaviors, and sensory function; eTable1 provides details regarding variable coding and question content of each exposure.

#### Cognitive tests

The psychometric properties and factor structure of the cognitive test battery in LASI-DAD wave 1 has been described previously.^(13)^ The battery, shown in Figure 1, has tests of memory, executive functioning and attention, language, orientation to place and time, and visuospatial functioning. Tests were selected to be comparable to the Harmonized Cognitive Assessment Protocol (HCAP) in other countries and adapted to the Indian context, given issues related to culture, language, and literacy.^(7,13)^ Some improvements in tests, administration, and scoring rules were introduced in wave 2, thus testing across waves was not identical. For example, administration of Symbol Cancellation shifted from paper in wave 1 to a tablet version in wave 2. To keep the symbols the same size given a smaller tablet screen, there were fewer symbols in wave 2. As an example of shifts in scoring rules, to the question "What do you do with a hammer," in wave 1 the correct answer was to pound a nail into something. In wave 2, we allowed responses of “to pound something” as correct, resulting in remarkably improved performance in wave 2. Adjustments made to the cognitive tests for wave 2 are in eTable2.

**Figure 1.**
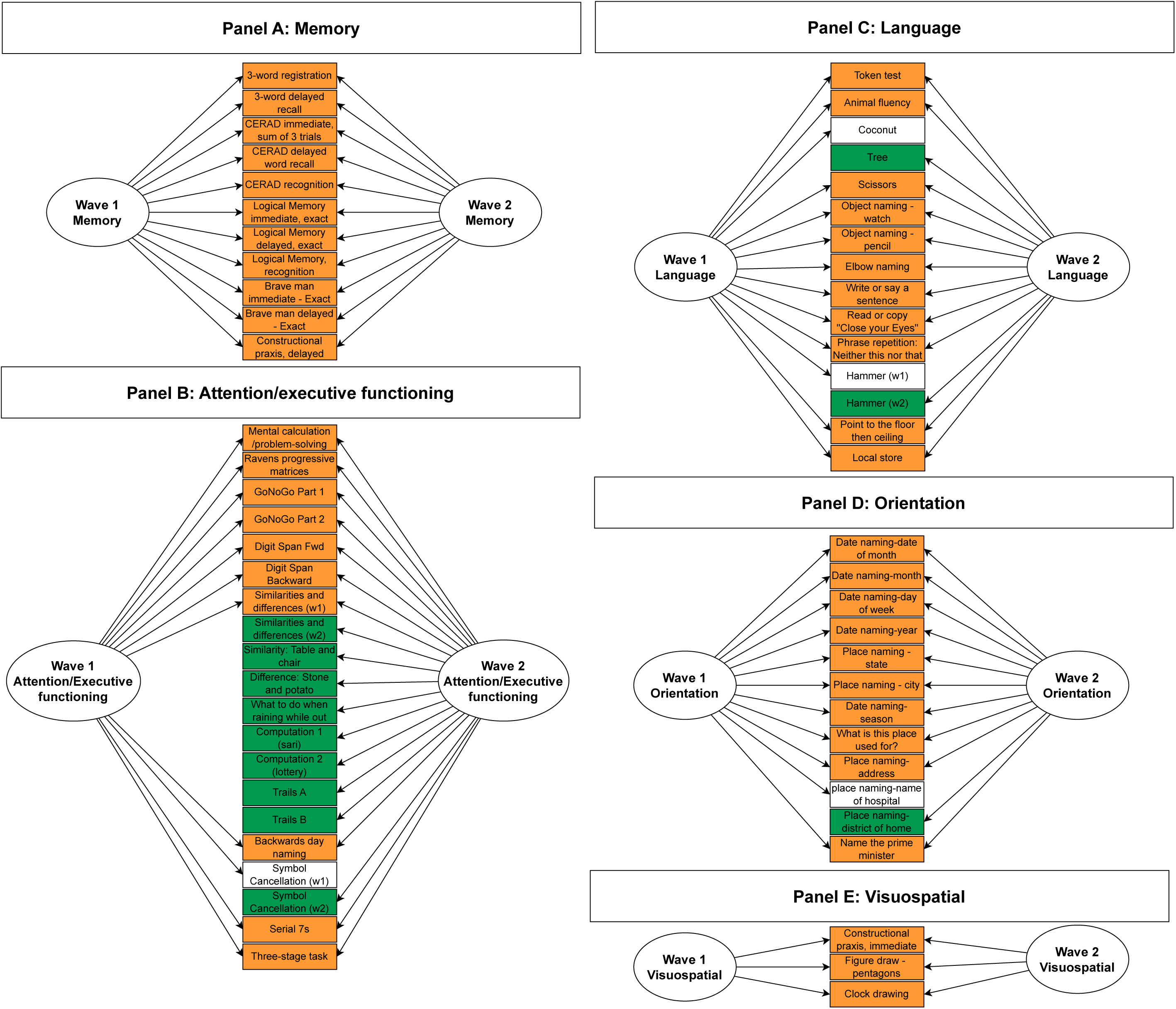
Structural equation model diagrams show cognitive tests in each cognitive domain: Results from LASI-DAD (N=6,168) Legend. Unobserved or latent variables, shown as circles, represent cognitive performance at waves 1 and 2 and reflect observed cognitive test scores, shown in rectangles. The general cognitive outcome was comprised of all available tests across all cognitive domains, and is not explicitly shown in this Figure. Colors for observed test scores are used to show overlap of test items across waves: orange signifies the cognitive test item was administered the same way in both waves; white signifies the item was in wave 1 only, and green signifies the item was in wave 2 only. Not shown in this figure are residual correlations to capture bifactor structures (see supplemental materials) or temporal correlations between the same tests administered over time (tests in orange in this Figure). LASI-DAD: Longitudinal Aging Study in India – Diagnostic Assessment of Dementia; CERAD: Consortium to Establish a Registry for Alzheimer’s Disease

#### Demographic characteristics

We included variables for age, sex, education (any formal schooling vs none), marital status (married or living with a partner, vs not), caste (scheduled caste or tribe versus none or Other Backwards Caste),^(14)^ height (meters), weight (kilograms), body mass index (considered separately from its components of height and weight), number of living children, mother’s and father’s education, and current work status (working, not working at the time of the LASI core survey).

#### Health and everyday function

Cardiovascular characteristics included self-reported history of a health care professional’s diagnosis of high blood pressure, any heart problem, stroke, heart attack, diabetes, and high cholesterol. Additional health factors included self-reported injurious falls and self-rated health. We also calculated summary measures of everyday functioning from indicators of difficulty in performing any of six basic activities of daily living (ADL) and in instrumental ADLs (IADL) (eTable1).^(15)^ We created a summary score for sleep problems based on the Jenkins Sleep Scale.^(16)^

#### Objective physical activity

The core LASI interview included a standard balance test^(17)^ and a 4-meter gait speed task.

#### Health behaviors

The LASI Core study asked about past-week frequency spent doing vigorous and moderate activities, current smoking status (reference: never and former smokers), and any alcohol use in the past 3 months.

#### Sensory function

We considered self-report of a health care professional’s assessment of any hearing or ear-related condition, as well as objectively measured moderate or severe impairment in near or distance vision based on vision acuity testing, with correction as needed.^(18)^

### Analysis plan

We characterized variables in the LASI-DAD sample using appropriate descriptive statistics. The primary analysis was conducted in two steps. First, in a series of confirmatory factor analyses (CFA) we summarized cognitive test information into cognitive domains, largely replicating the factor structure of cognitive data from LASI-DAD wave 1,^(13)^ and cocalibrated cognition across the waves to predict comparable factor scores for general and domain-specific cognition (memory, executive function/attention, language/fluency, orientation, visuospatial function) at each wave (see Supplemental Methods and Results). Next, among the N=2565 participants seen in both LASI-DAD waves 1 and 2, we evaluated associations of 33 risk and protective factors with rates of cognitive change using mixed effects models with random effects for people and time.^(19)^ In these mixed effects models, cognitive outcomes at waves 1 and 2 were regressed on indicators for time, a risk factor, and their interaction; the interaction of time with a risk factor provides an estimate of its effect on annualized cognitive change. Time since baseline was modeled as a continuous variable, such that slopes from models can be interpreted as annualized rates of change. To assist with interpretability of findings, we divided coefficients by one tenth of the age coefficient for cognitive decline to scale associations per year of older age. All models were adjusted for age, sex, and schooling, and their interactions with time.

Parametric analyses were conducted using Mplus, version 8.7.^(20)^

## Results

The LASI-DAD sample includes N=6168 participants, of whom N=2566 were in both waves; there were N=4096 respondents in wave 1 and N=4638 in wave 2 (Table 1). LASI-DAD wave 2 took place between 2.8 and 6.4 years after wave 1 (median 4.5 years). At wave 1, among those with longitudinal follow-up the mean age of the sample was 68.3 (SD=6.5) years (range 60, 105), 56.5% of participants were female, 51.9% had formal schooling, 63% were married or cohabitating with a partner, and 24% were from a scheduled caste or tribe. At the time of the LASI core interview, participants reported having 4 children on average, and 34% of the sample reported they were currently working.

### Average rate of cognitive change

Among N=2566 participants with longitudinal follow-up, the mean rate of general cognitive change was -0.03 standard deviations (SD) per year in the full sample (Table 2). Rate of change was progressively steeper with older age: -0.021 SD/year among 60-69 year olds, -0.039 SD/year among 70-79 year olds, and -0.075 SD/year among participants aged 80+ years. We observed analogous patterns of progressive decline for memory, executive function/attention, visuospatial functioning, and orientation. Language showed improvement over time (0.027 SD/year) on average, with decline apparent only among participants aged 80+ years at baseline (-0.030 SD/year).

### Predictors of cognitive change

We tested correlates of change in general cognitive performance using exposures from the LASI Core survey (Table 3). Among demographic characteristics, older age, any education, heavier weight, and greater body mass index were associated with steeper cognitive decline while being coupled (married or living with a partner), having more living children, and working at the time of the LASI Core survey were associated with less steep general cognitive decline. Across all risk and protective factors, education had the strongest association with cognitive decline, followed by parental education, but in unexpected directions; this association persisted after treating education continuously and in groups stratified by baseline age. Among health history variables, most self-reported cardiovascular characteristics (e.g., any heart problem, high blood pressure, high cholesterol, heart attack, and diabetes), were associated with subsequent steeper cognitive decline. When rescaled by dividing by a tenth of the age-decade coefficient for cognitive decline, these associations were comparable to 7-12 years of older age. Among self-reported health and functioning characteristics, report of any ADL difficulty and more sleep problems were associated with steeper general cognitive decline; these associations were each comparable to about 5 years of older age. Among sensory functions, neither self-reported hearing nor objectively measured vision were associated with changes in general cognitive performance. eTables 5-9 show associations with cognitive change using specific cognitive domains; coefficients were in consistent directions for nearly all domains for each risk factor.

## Discussion

We replicated the factor structure of cognition from the first wave of LASI-DAD in India and extended it to a newly collected second wave. We then evaluated associations of a variety of demographic and health risk and protective factors with cognitive change using up to 6.4 years of longitudinal follow-up in this nationally representative population. Most risk factors were associated with changes in general cognitive performance in the same direction as previous findings from high-income contexts, with exceptions of education and smoking. Expected relationships were strongest for self-reported history of a health care professional’s diagnosis of cardiovascular conditions. The longitudinally calibrated summary cognitive factor scores for general and domain-specific cognition reported here can be used for future substantive research. Psychometric evaluation of measurement invariance suggests it is unlikely that differences in administration, scoring, or mode of test administration are responsible for cognitive changes we observed, and thus temporal changes in these factors can be attributed to changes in cognitive performance.

The 2024 Lancet Commission on dementia released an update to their 2020 report, which suggested that up to 40% of the risk of dementia may be attributable to 14 potentially modifiable risk factors for dementia: less education, hearing loss, traumatic brain injury, hypertension, excessive alcohol consumption, obesity, smoking, depression, social isolation, physical inactivity, air pollution, diabetes, high LDL cholesterol, and untreated vision impairment. Among the risk and protective factors tested in this study, we evaluated 12 of the 14 risk factors, or proxies of them, based on data from the core LASI survey. We did not assess social isolation or air pollution, although such data are available for future research. While the LASI survey did not ask about traumatic brain injury in midlife, we did consider injurious falls within 2 years of the LASI survey. Of note, the LASI-DAD survey asked about traumatic brain injury, but we restricted ourselves to exposures collected from the LASI Core survey to maintain temporality between exposures and cognitive change. Falls are the most common cause of traumatic brain injury among older adults,^(21)^ and while not statistically significantly associated with cognitive change, the association is in the expected direction. Additionally, while the Lancet Commission specifies high LDL cholesterol, we used a variable for physician’s report of any high cholesterol. Thus, for the 12 risk factors available in our study, we found that with exceptions of smoking and education, these factors increased rate of cognitive decline among older Indian adults over the course of up to 6.4 years.

Although this paper focuses on predictors of cognitive decline and not dementia per se, cognitive decline is a key feature of dementia. Future studies are needed to evaluate whether the same risk and protective factors here are also associated with risk of dementia and their population attributable fractions for India and other low- and middle-income countries. An important feature of population attributable fractions, used in the 2024 Lancet Commission report, is that they are a function of the prevalence of a risk factor with dementia, as well as strength of the association.^(22)^ Both prevalence and strength vary across social factors and geography, such that the potential reduction in prevalence may even be greater in low- and middle-income countries such as India due to greater prevalences of most risk factors.^(23)^ One exception would be alcohol consumption, which is less common in India^(22)^ and in our study not associated with cognitive change.

An unexpected finding is that more education was associated with steeper cognitive decline. There are several potential explanations for this finding. One explanation might be survival bias: more educated people could receive better medical treatment for cardiovascular disease and live longer with risk factors than those with less or no formal education.^(24)^ Mortality was higher among LASI-DAD participants with lower levels of education, and general cognitive performance in wave 1 was higher among those who survived to wave 2 than among those who did not return (Table 1); thus, decline may be masked in the lower-education group.^(29)^ Or, perhaps education conditions people to take tests, which is why it is normally recommended to adjust for years of education on most neuropsychological tests.^(25–27)^ However, half of LASI-DAD reported little to no formal schooling, which is average for this nationwide sample in India. Thus, these individuals may be more likely to benefit from repeat exposure to cognitive testing.

Another explanation is that according to cognitive reserve theory, once a certain pathological threshold is met (i.e., degree of cortical atrophy) individuals with higher education show a steeper rate of decline because they previously had been able to maintain higher than expected performance in the face of pathology.^(28)^ While LASI-DAD is not a study of patients with dementia, this may be a partial explanation. It is possible that each of these explanations play a role and further research, beyond the scope of this paper, is needed to disentangle them.

Advantages of this study include the large, nationally representative sample of older adults in a low-to-middle income country and the detailed battery of cognitive tests available in LASI-DAD. Cognitive tests were selected to be comparable to other HCAP studies but were also tailored as necessary to the Indian context.^(7)^ Correlations among cognitive tests in the HCAP battery are well-described by factors for memory, executive functioning, language, visuospatial functioning, and orientation, as was true of the first wave of LASI-DAD.^(13)^ Longitudinally calibrated domain scores developed in this study are optimized for the evaluation of both cognitive levels and cognitive decline. The scores are purified of nuisance differences in the HCAP battery by accounting for residual correlations among some indicators via bifactor structures.

Nevertheless, this study is not without limitations. Foremost, we had just two waves separated by up to six years of follow-up to evaluate changes in cognition. Additional follow-up would serve to strengthen this study. A major event intervening waves 1 and 2 was the global COVID19 pandemic and its associated lockdowns, which had a profound impact on daily life and mortality especially in India.^(30)^ Because of this, observed rates of cognitive change should be interpreted carefully and may not generalize to other contexts or time periods. Another limitation is that most of the risk and protective factors we evaluated were based on self-report from participants. Especially for health history characteristics, asking whether a doctor or medical professional has ever told a participant they have a certain condition can be biased given unequal access to health care in India. Self-reported variables about cardiovascular risk factors were among the strongest correlates of cognitive change in LASI-DAD, potentially because of an over-representation of severe cases due to challenges in accessing the healthcare system.

We longitudinally calibrated general and domain-specific performance in LASI-DAD, a large, population representative sample of older adults in India. We evaluated risk and protective factors for cognitive change, including most risk factors established by the 2024 Lancet Commission on dementia, with results suggesting that these risk factors, identified using evidence largely from high-income countries, are also related to cognitive decline in India. The single risk factor models we estimated serve an important purpose in identifying potentially relevant predictors, simplifying interpretation by and providing estimates of total effects that may be informative in their own right. However, given that many of the risk factors are likely to be correlated, these models conflate the effect of one factor with that of others. For instance, the estimated effect of BMI may partially capture the downstream influence of cardiovascular conditions, or vice versa. Future research is needed to disentangle total effects from indirect effects.

## Supporting information

Manuscript tables

## Data Availability

Data and codebooks for LASI Core and LASI-DAD are publicly available at https://g2aging.org/auth/signin?next=/lasi/download. Data are shared via this website with a signed data access agreement.

## Acknowledgements

We thank the participants and families who participated in the LASI-DAD study, the staff at the study sites, and the personnel involved in the data collection and data release.

We obtained ethics approval from the Indian Council of Medical Research (2202-16741/F1) and all collaborating institutions, including the University of Southern California (UP-15-00684), All India Institute of Medical Sciences, New Delhi, Venu Geriatric Center, New Delhi, All India Institute of Medical Sciences, Bhubaneshwar, All India Institute of Medical Sciences, Rishikesh, Uttarakhand, Government Medical College, Chandigarh, Punjab, Aster MIMS Kannur, Kerala, Grants Medical College & JJ Hospital, Mumbai, Guwahati Medical College, Guwahati, All India Institute of Medical Sciences, BHU, Varanasi, Jawaharlal Institute of Postgraduate Medical Education and Research, Puducherry, Medical College, Kolkata, National Institute of Mental Health and Neurosciences, Bengaluru, St. John’s Medical College, Bengaluru, All India Institute of Medical Sciences, Bibinagar, Hyderabad, Sher-e-Kashmir Institute of Medical Sciences, Srinagar, All India Institute of Medical Sciences, Mangalagiri, Andhra Pradesh, All India Institute of Medical Sciences, Bhopal Madhya Pradesh, and All India Institute of Medical Sciences, Raipur.

## Contributor’s statement

ALG designed the study, led the study’s analytic strategy, and wrote the first draft of the manuscript. ALG, EN, MAR, PYK, EM, MG, EMC, SD, ABD, and JL contributed to the conceptualization of the study. ALG, EN, PYK, and EM were responsible for data curation. ALG and EN conducted formal analyses. EMC, SD, ABD, and JL obtained funding for the study. ALG, EN, MAR, EM contributed to the methods used in the manuscript. SD, PYK, and JL led the project’s administration. ALG contributed to the data visualization. All authors have accessed and verified all the included data. All authors reviewed and edited the paper for important intellectual content. ALG had final responsibility for the decision to submit to publication.

