## Supplementary material for "Risk and protective factors against cognitive decline in older adults from a nationally representative sample in India: Results from the LASI-DAD": Manuscript tables

Table 1. Descriptive characteristics of the LASI-DAD sample (N=6,168)

| Characteristic | Waves 1 and 2 (N=2566) | Wave 1 only (N=1530) | Wave 2 only (N=2072) |
| --- | --- | --- | --- |
|  | Mean (SD) or N (%) | Mean (SD) or N (%) | Mean (SD) or N (%) |
| Demographic |  |  |  |
| Age at LASI-DAD wave 1, mean (SD) | 68.3 (6.5) | 72.1 (8.7) | 65.6 (8.5) |
| Female sex, n (%) | 1450 (56.5) | 757 (49.5) | 1242 (59.9) |
| Any formal schooling, n (%) | 1332 (51.9) | 765 (50.0) | 706 (34.1) |
| Married, n (%) | 1624 (63.3) | 839 (54.8) | 1165 (56.2) |
| Scheduled caste or tribe, n (%) | 615 (24.0) | 341 (22.3) | 585 (28.2) |
| Height, meters, mean (SD) | 1.5 (0.1) | 1.5 (0.1) | 1.5 (0.1) |
| Weight, kilograms, mean (SD) | 54.6 (12.9) | 53.0 (13.0) | 52.6 (12.6) |
| Body mass index, kilograms/meters2, mean (SD) | 22.7 (4.8) | 22.2 (4.9) | 22.1 (4.7) |
| Number of living children, mean (SD) | 3.9 (1.8) | 3.9 (1.9) | 3.7 (1.8) |
| Mother's education, any, n (%) | 47 (1.9) | 34 (2.4) | 34 (1.7) |
| Father's education, any, n (%) | 207 (8.5) | 130 (9.2) | 118 (6.0) |
| Currently working, n (%) | 874 (34.1) | 372 (24.5) | 836 (40.4) |
| Health history |  |  |  |
| High blood pressure, n (%) | 978 (38.1) | 627 (41.4) | 649 (31.4) |
| Diabetes, n (%) | 387 (15.1) | 332 (21.9) | 255 (12.3) |
| Any heart problem, n (%) | 168 (6.5) | 109 (7.2) | 66 (3.2) |
| Stroke, ever, n (%) | 56 (2.2) | 64 (4.2) | 48 (2.3) |
| High cholesterol, n (%) | 123 (4.8) | 67 (4.4) | 76 (3.7) |
| Heart attack, n (%) | 88 (3.4) | 63 (4.2) | 31 (1.5) |
| Injurious fall, n (%) | 431 (71.5) | 218 (66.1) | 326 (68.9) |
| Self-reported health and function |  |  |  |
| Any ADL difficulty, N (%) | 507 (19.8) | 444 (29.4) | 435 (21.1) |
| IADL difficulty (sum of 16 indicators), mean (SD) | 4.7 (4.2) | 6.2 (4.9) | 5.2 (4.6) |
| Sleep problems (higher is worse), mean (SD) | 0.4 (0.5) | 0.4 (0.5) | 0.4 (0.5) |
| Self-rated health, excellent or good, n (%) | 1282 (50.4) | 650 (44.2) | 1080 (52.9) |
| CIDI major depression, n (%) | 178 (7.0) | 104 (7.1) | 178 (8.8) |
| Objective physical activity |  |  |  |
| Balance test, can do both semi and full stand, n (%) | 1751 (76.4) | 794 (64.5) | 1342 (74.6) |
| Seconds to walk 4 meters, mean (SD) | 5.9 (1.9) | 6.6 (2.5) | 5.9 (1.9) |
| Health behaviors |  |  |  |
| Vigorous activity, at least a few times a week, n (%) | 605 (23.7) | 236 (15.7) | 532 (25.8) |
| Moderate activity, at least a few times a week, n (%) | 1464 (57.2) | 682 (45.3) | 1184 (57.5) |
| Any alcohol in last 3 months, n (%) | 184 (7.2) | 85 (5.6) | 170 (8.3) |
| Current smoker, n (%) | 385 (15.1) | 235 (15.6) | 312 (15.2) |
| Sensory function |  |  |  |
| Any hearing/ear-related condition, n (%) | 246 (9.6) | 192 (12.7) | 195 (9.4) |
| Near vision impairment in better eye, n (%) | 1639 (68.1) | 978 (71.2) | 1309 (69.9) |
| Distance vision impairment in better eye, n (%) | 757 (31.4) | 581 (42.2) | 630 (33.6) |

Legend. SD: standard deviation; ADL: activities of daily living; IADL: instrumental activities of daily living; CIDI: Composite International Diagnostic Interview.

Table 2. Annual rates of cognitive change in factor scores overall and by age: Results from LASI-DAD (N=2,566)

|  | Full sample | |  | Age 60-69 years | |  | Age 70-79 years | |  | Age 80+ years | |
| --- | --- | --- | --- | --- | --- | --- | --- | --- | --- | --- | --- |
| Cognitive domain | Estimate | Z |  | Estimate | Z |  | Estimate | Z |  | Estimate | Z |
| General cognitive performance | -0.029 | -11.2 |  | -0.021 | -6.6 |  | -0.039 | -7.8 |  | -0.075 | -6.8 |
| Memory | -0.026 | -8.2 |  | -0.017 | -4.4 |  | -0.039 | -6.5 |  | -0.061 | -4.8 |
| Executive function/attention | -0.034 | -12.8 |  | -0.027 | -8.6 |  | -0.040 | -7.9 |  | -0.074 | -6.6 |
| Language/fluency | 0.027 | 7.6 |  | 0.038 | 9.1 |  | 0.013 | 1.9 |  | -0.030 | -2.0 |
| Orientation | -0.015 | -5.6 |  | -0.009 | -2.8 |  | -0.019 | -3.8 |  | -0.060 | -4.9 |
| Visuospatial | -0.038 | -13.3 |  | -0.033 | -9.5 |  | -0.049 | -9.0 |  | -0.039 | -3.4 |

Legend. Rates of cognitive change, which are coefficients for time from random effects models, are shown on a metric in which cognitive outcomes are standardized to a N(0,1) distribution, and are thus scaled to a Cohen’s d. Negative estimates suggest cognitive decline. Columns of Z-scores show the quotient of the modeled estimate and the standard error of the estimate.

Table 3. Associations of risk factors for dementia with change in general cognitive performance: Results from LASI-DAD (N=2,566)

| Variable | Change in general cognitive performance | | | |
| --- | --- | --- | --- | --- |
|  | Beta | SE | z-statistic | Age- standardized beta |
| Demographics and family background | | | |  |
| Age, per 10 years | -0.019 | 0.004 | -4.60 | 10.00 |
| Female sex | -0.014 | 0.006 | -2.49 | 7.37 |
| Any formal schooling | -0.052 | 0.005 | -9.98 | 27.37 |
| Married | 0.014 | 0.006 | 2.41 | -7.37 |
| Scheduled caste or tribe | -0.004 | 0.006 | -0.67 | 1.82 |
| Height, meters | 0.004 | 0.039 | 0.11 | -1.74 |
| Weight, kilograms | -0.001 | 0.000 | -2.35 | 0.40 |
| Body mass index | -0.002 | 0.001 | -2.75 | 0.83 |
| Number of living children | 0.005 | 0.001 | 3.74 | -2.08 |
| Mother's education, any | -0.034 | 0.016 | -2.12 | 15.45 |
| Father's education, any | -0.028 | 0.009 | -3.01 | 12.17 |
| Currently working | 0.008 | 0.005 | 1.40 | -3.81 |
| Health history |  |  |  |  |
| High blood pressure | -0.014 | 0.005 | -2.75 | 6.67 |
| Diabetes | -0.021 | 0.007 | -3.22 | 9.55 |
| Any heart problem | -0.020 | 0.009 | -2.24 | 9.09 |
| Stroke | -0.016 | 0.016 | -1.02 | 7.27 |
| High cholesterol | -0.024 | 0.009 | -2.58 | 10.91 |
| Heart attack | -0.026 | 0.012 | -2.19 | 11.82 |
| Injurious fall | -0.012 | 0.013 | -0.92 | 7.06 |
| Self-reported health and function | | |  |  |
| Any ADL difficulty | -0.010 | 0.007 | -1.50 | 4.76 |
| IADL difficulty | 0.000 | 0.001 | 0.08 | 0.00 |
| Sleep problems | 0.011 | 0.005 | 2.17 | -5.00 |
| Self-rated health (Excellent or good) | 0.007 | 0.005 | 1.53 | -3.04 |
| Probable major depression (CIDI) | 0.019 | 0.010 | 1.94 | -8.26 |
| Objective physical activity | |  |  |  |
| Balance test | 0.011 | 0.006 | 1.70 | -5.24 |
| Average walking speed (seconds) | 0.000 | 0.001 | -0.23 | 0.00 |
| Health behaviors |  |  |  |  |
| Vigorous activity | 0.004 | 0.006 | 0.76 | -1.82 |
| Moderate activity | -0.005 | 0.005 | -1.06 | 2.17 |
| Alcohol use, past 3 months | 0.012 | 0.010 | 1.18 | -5.45 |
| Currently smokes | 0.022 | 0.007 | 2.98 | -10.48 |
| Sensory function |  |  |  |  |
| Any hearing/ear-related condition | 0.016 | 0.009 | 1.83 | -6.96 |
| Near vision impairment in better eye | 0.003 | 0.005 | 0.57 | -1.25 |
| Distance vision impairment in better eye | 0.006 | 0.006 | 1.13 | -2.40 |

Legend. Shown are regressions of general cognitive change on risk factors. Each row is from a separate model. All models are adjusted for age, sex, and education. The "Beta" column shows the association of each exposure with change in cognition; negative values imply the predictor is associated with steeper cognitive decline. The "z-statistic" column is the beta/SE. The column "age-standardized beta" shows the coefficient divided by the age coefficient from that model, which reflects the association on a standardized metric with respect to age. SE: standard error; ADL: activities of daily living; IADL: instrumental activities of daily living; CIDI: Composite International Diagnostic Interview.
