## Supplemental materials for "Risk and protective factors against cognitive decline in older adults from a nationally representative sample in India: Results from the LASI-DAD"

Supplemental Methods and Results: Longitudinal cocalibration of cognition in LASI-DAD

**Methods**. We fit unidimensional CFA models to cognitive test items separately for general cognitive performance and each domain (memory, attention/executive functioning, language, and orientation), and for each wave. In CFA, latent variables are postulated to represent shared, or common, covariation among observed indicators in the model. Because of this, factors representing the latent variables are said to filter out measurement error, or residual variance from tests not shared with other tests.^(1)^ As necessary, we estimated residual correlations among items, using a bifactor approach,^(2)^ to improve fit of CFA models to the data if models did not fit well initially. After fitting models for each domain and identifying satisfactory model fit, we estimated a model for general cognitive performance based on all available cognitive test items.

Once we did this for cognitive data in wave 1, we applied the same models to wave 2 and evaluated fit. After finding optimal-fitting solutions for each cognitive domain and general cognitive performance at each wave, we modeled wave 1 and wave 2 cognitive performance together using a CFA model with separate (but correlated) factors for each wave, bifactors for methods correlations, and cross-wave correlations between indicators common between the waves. To derive factor scores from CFAs, we used a model with a robust maximum likelihood estimator from which factor scores were estimated based on expected a posteriori methods. To evaluate fit of CFA models, we considered the root mean squared error of approximation (RMSEA; <=0.05 is excellent), comparative fit index (CFI; >0.95 is excellent), and standardized root mean squared residual (SRMR; <=0.08 is excellent).^(3)^ The latent variable approach used here has advantages over other methods of creating summary factors. The approach is better than relying only on individual tests in common between waves of LASI-DAD because new tests in wave 2, as well as discontinued tests from wave 1, still provide some information about a participant's cognitive ability.^(4-6)^ A distinct advantage of latent variable methods over alternative approaches, such as standardizing raw test scores and averaging them together, is that given changing cognitive tests across waves in LASI-DAD (eTable 2), cognitive test item parameters are freely estimated but fixed to be the same across waves. These cross-wave constraints on item parameters ensure that cognitive summary factors from wave 1 are psychometrically comparable to cognitive summary factors from wave 2, and moreover that the factors are flexibly estimated to reflect the data.

**Results**. Unidimensional CFAs of domains for general cognitive performance, memory, executive functioning/attention, language/fluency, visuospatial ability, and orientation each fit well to the data separately in waves 1 and 2, and in a combined two-factor CFA model across all waves (eTable 3). eFigure 1 displays histograms by wave of general and domain-specific cognitive factor scores estimated from the models. eTable 4 provides model-estimated factor loadings for each cognitive test indicator from the longitudinal CFA models for each cognitive domain. All standardized factor loadings, bounded between -1 and 1, ranged between 0.4 and 0.9, suggesting an acceptable degree of shared covariance among the cognitive test items. Consistent with unidimensionality of each cognitive domain, the relative magnitudes of standardized loadings within a domain are fairly similar. Marginal reliabilities of the scores are high across the range of each latent cognitive trait, with the exceptions of language and orientation at higher levels of ability (eFigure 2).

We tested levels of temporal measurement invariance of the factor structure of the general cognitive performance factor. Absolute fit was excellent for multiple-group models conforming to configural (RMSEA: 0.04; CFI: 0.95; SRMR: 0.06), metric (RMSEA: 0.04; CFI: 0.96; SRMR: 0.07), and scalar invariance (RMSEA: 0.04; CFI: 0.94; SRMR: 0.07). These fit statistics suggest the covariance among cognitive tests in waves 1 and 2 was similar. Moreover, because these tests allow the mean and variance of the cognitive latent variable to differ by LASI-DAD wave, evidence for scalar invariance suggests it is unlikely that differences in administration, scoring, or mode are responsible for changes in the general factor and thus temporal changes in the factor can be attributable to changes in cognitive performance.

**Imputation**. Missing data can have a detrimental effect on surveys when not handled well. This is particularly true for the large cognitive battery and informant scales in LASI-DAD, because a small fraction of missings at the item level can lead to a large fraction missing at higher levels of aggregation. For example, only 4% of the items in the Hindi Mental State Exam (HMSE) were missing in LASI-DAD wave 1, but 27% of the participants had at least one item missing, which would lead to the whole scale being missing. For the Jorm IQCODE, these numbers were 9% and 50%, respectively.

Thus, we imputed missing data in LASI-DAD. The imputation methods have been summarized in the supplementary material to Gross et al.^(1)^ and described in more detail in Wilkens et al.^(2)^. The methods were inspired by the cognition imputation methods used in the Health and Retirement Study (HRS)^(3)^. Briefly, binary, ordinal, and unordered categorical variables were imputed using binary, ordered, and multinomial logistic regression models, respectively. Count variables such as animal naming were imputed using a negative binomial regression model. To correctly reflect the correlations among the variables, a chained imputation procedure was used, ^(4,5)^ in which each variable is imputed sequentially, including the (potentially imputed) other variables as covariates, and iteratively cycling through the variables, allowing updated imputations of a variable to be used as covariates in the next iteration of imputations of other variables. However, because the large number of variables would lead to numerical problems and overfitting when used as covariates as-is, we used aggregates instead of raw variables as covariates. These aggregates were inspired by the factor model as described above and used in Gross et al. (2020), although instead of estimating the full factor model within each imputation step, which would have been computationally prohibitive, we used simple unweighted or weighted sum scores. For example, for imputing the city naming item, which is part of orientation to place, the other orientation to place items (e.g., state, address) were used in raw form, but orientation to time was included as a simple sum score (0-5), and memory was included as a weighted aggregate of 84 items. For, say, immediate 10-word recall, orientation would be included as a single aggregate, but memory would be broken down into delayed memory, recognition memory (i.e., two narrow domain aggregates), and several immediate memory variables and sum scores. In addition to the (other) cognition and informant variables, demographics, socio-economic variables, and health/disability variables (including mental health), as well as cognition variables from the LASI core survey were included as covariates in all models. If necessary, these were themselves imputed in a preliminary step before imputing the cognition and informant report variables.

Wave 2 introduced a few new considerations. The Trail Making Test was added, and one set of variables from this consists of the average times per path, which are continuous variables. We imputed these (after log transformation to reduce skewness) using predictive mean matching,^(6)^ which is a semi-parametric method that is more robust to model misspecification than parametric methods,^(7)^ which is especially salient for continuous variables. Because of the very different nature of these variables, we imputed these as a small chained imputation (i.e., just the three Trail Making average time variables) after imputing all the other variables.

The main new consideration for wave 2 was whether and how to use the longitudinal dimension. In addition to the (other) wave 2 variables and the demographics and other covariates from the core data, all LASI-DAD wave 1 variables are available as potential covariates. Because this potentially exacerbates the risks of numerical problems and overfitting, we considered a wide range of modeling options, with inclusion or exclusion of different types of variables (health, economic variables) or different waves (LASI-DAD wave 1, LASI Core wave 1). To select the preferred model, we used Akaike's Information Criterion (AIC),^(8)^ a commonly used measure for assessing trade-offs between fit and parsimony. The AIC was designed to select the model with the best out-of-sample prediction. Because it is undesirable to select widely different models for different variables within the same imputation chain (e.g., which covariates are used affects the sample composition: LASI-DAD wave 1 variables are unavailable for respondents who did not participate in LASI-DAD wave 1), we computed the sum of the AICs for the different variables and selected the model specification that minimized this sum among the variations considered. We ensured that the different models that were being compared used the same sample. The most surprising result was that the LASI-DAD wave 1 variables were not included, but the LASI Core wave 1 variables were. This does not mean that there is no serial correlation in the variables in LASI-DAD (there is), but most of this serial correlation is catered for by the contemporaneous (LASI-DAD wave 2) or earliest (LASI Core wave 1) analogous measures. Adding these variables from LASI-DAD wave 1 added a large number of parameters to the model while only modestly improving the fit. Hence, the model with these variables included would lead to noisier imputations than with them excluded.

Supplemental References regarding imputation

1. Gross AL, Khobragade PY, Meijer E, Saxton JA. Measurement and Structure of Cognition in the Longitudinal Aging Study in India-Diagnostic Assessment of Dementia. J Am Geriatr Soc. 2020;68 Suppl 3(Suppl 3):S11-S19.
2. Wilkens J et al. (2024). Harmonized LASI-DAD documentation, version A.4. Los Angeles, CA: Gateway to Global Aging Data.
3. Fisher GG, Hassan H, Faul JD, Rodgers WL, Weir DR (2017). Health and Retirement Study: Imputation of Cognitive Functioning Measures: 1992 - 2014 (Final Release Version): Data Description. Ann Arbor, MI: University of Michigan, Survey Research Center.
4. Raghunathan TE, Lepkowski JM, van Hoewyk J, Solenberger P. A multivariate technique for multiply imputing missing values using a sequence of regression models. Survey Methodology 2001; 27: 85-95.
5. Van Buuren S, Brand JPL, Groothuis-Oudshoorn CGM, Rubin DB. Fully conditional specification in multivariate imputation. Journal of Statistical Computation and Simulation 2006; 76: 1049–1064.
6. Little RJA. Missing-data adjustments in large surveys. Journal of Business & Economic Statistics 1988; 6: 287–296.
7. Kleinke K. Multiple Imputation Under Violated Distributional Assumptions: A Systematic Evaluation of the Assumed Robustness of Predictive Mean Matching. Journal of Educational and Behavioral Statistics 2017; 42: 371–404.
8. Amemiya T. Selection of regressors. International Economic Review 1980; 21: 331-354.

eTable 1. Details regarding variable coding and question content of each exposure measured in the LASI Core survey

| Exposure | Relevant details of variable coding or question content |
| --- | --- |
| Demographic |  |
| Age at LASI-DAD wave 1 | Self-evident |
| Female sex | Self-evident |
| Any formal schooling | Binary indicator for any formal schooling (reference group is none) |
| Married | Binary indicator for being married or cohabitating with someone else, versus not |
| Scheduled caste or tribe | Binary indicator for scheduled caste or tribe, versus other backward class or no caste |
| Height, meters | Winsorized the top 0.1% of values |
| Weight, kilograms | Winsorized the top 0.1% of values |
| Body mass index | kilograms/meters^2^ |
| Number of living children | Total number of living children, winsorized the top 1% of values, yielding a maximum of 9 children |
| Mother's education, any | Self-report of whether mother had any formal schooling |
| Father's education, any | Self-report of whether father had any formal schooling |
| Currently working | Binary indicator for whether participant self-reported currently working during the LASI core interview |
| Health history |  |
| High blood pressure | Self-evident |
| Diabetes | Self-evident |
| Any heart problem | Self-evident |
| Stroke, ever | Self-evident |
| High cholesterol | Self-evident |
| Heart attack | Self-evident |
| Injurious fall, past 2 years | Self-reported injurious fall within 2 years of the LASI core survey |
| Self-reported health and function |  |
| Any ADL difficulty | 6 ADLs included difficulty walking across a room, dressing, bathing/showering, eating, getting in or out of bed, toileting |
| IADL difficulty (sum of 16 indicators) | 16 IADLs included difficulty using telephone, taking medications, managing money, shopping for groceries, preparing hot meal, getting around, doing work around, walking 100 yards, sitting for 2 hours, getting up from a chair, clmbing 1 flight of stairs, Stooping/kneeling/crouching, Lift/carry 5 kilos, picking up a coin, reach with arms up, and push/pull large object |
| Sleep problems (higher is worse) | Average score of trouble falling asleep, waking up during the night, waking up too early, and feeling unrested during the day |
| Self-rated health, excellent or good | Self-evident |
| CIDI major depression | Lay diagnostic interview, administered during LASI Core survey |
| Objective physical activity |  |
| Balance test, can do both semi and full stand | Binary indicator of whether participants successfully completed both semi and full stance, versus not |
| Seconds to walk 4 meters | Seconds to walk 4 meters at regular pace. |
| Health behaviors |  |
| Vigorous activity, at least a few times a week | Binary indicator of the self-reported frequency of vigorous activity, coded as everyday/more than once weekly, versus once a week or less. Examples of vigorous activities include running, swimming, going to a health center or gym, cycling, digging with a spade or shovel, heavy lifting, chopping, farm work, fast bicycling, or cycling with loads. |
| Moderate activity, at least a few times a week | Binary indicator of the frequency of moderate activity, coded as everyday/more than once weekly, versus once a week or less. Examples of moderate activities include house cleaning, handwashing clothes, fetching water or wood, drawing water from a well, gardening, bicycling at a regular pace, walking at a moderate pace, dancing, or floor or stretching exercises |
| Any alcohol in last 3 months | Binary indicator of self-reported alcohol use in the last 3 months |
| Current smoker | Binary indicator of self-reported current smoking, versus never or former smoking |
| Sensory function |  |
| Any hearing/ear-related condition | Binary indicator for self-report of a diagnosis of any hearing or ear-related condition |
| Near vision impairment in better eye | Binary indicator for objectively tested near vision, indicating moderate to severe vision impairment (blindness) versus no or mild visual impairment |
| Distance vision impairment in better eye | Binary indicator for objectively tested distance vision, indicating moderate to severe vision impairment (blindness) versus no or mild visual impairment |

Legend. ADL: activities of daily living; IADL: instrumental activities of daily living; CIDI: Composite International Diagnostic Interview

eTable 2. Test adaptations between LASI-DAD waves 1 and 2

| Cognitive test item | Wave of administration | Test adaptation |
| --- | --- | --- |
| **Memory** |  |  |
| 3-word registration | 1, 2 | No change |
| 3-word delayed recall | 1, 2 | No change |
| CERAD immediate recall, sum of 3 trials | 1, 2 | No change |
| CERAD delayed word recall | 1, 2 | No change |
| CERAD recognition | 1, 2 | No change |
| Logical Memory immediate, exact | 1, 2 | No change |
| Logical Memory delayed, exact | 1, 2 | No change |
| Logical Memory, recognition | 1, 2 | No change |
| Brave man immediate - Exact | 1, 2 | No change |
| Brave man delayed - Exact | 1, 2 | No change |
| Constructional praxis, delayed | 1, 2 | No change |
| **Executive function/attention** |  |  |
| Trails A | 2 | Newly added in Wave 2 |
| Trails B | 2 | Newly added in Wave 2 |
| GoNoGo Part 1 | 1, 2 | No change |
| GoNoGo Part 2 | 1, 2 | No change |
| Similarities and differences (w1) | 1 | Four items were asked about similarities (elephant and monkey; rose and jasmine) and differences (lie and mistake; river and pond) |
| Similarities and differences (w2) | 2 | In addition to the four items in wave 1, two additional items were added in wave 2 (similarity between table and chair; difference between stone and potatoe) |
| Digit Span Forward | 1, 2 | No change |
| Digit Span Backwards | 1, 2 | No change |
| Symbol Cancellation (w1) | 1 | In wave 2, this was administered via paper and pencil. |
| Symbol Cancellation (w2) | 2 | In wave 2, this was administered using a tablet. |
| Three-stage task | 1, 2 | No change |
| Serial 7s | 1, 2 | No change |
| Backwards day naming | 1, 2 | No change |
| What to do when raining while out | 2 | Newly added in Wave 2 |
| Computation 1 (sari) | 2 | Newly added in Wave 2 |
| Computation 2 (lottery) | 2 | Newly added in Wave 2 |
| Mental calculation /problem-solving | 1, 2 | No change |
| Ravens progressive matrices | 1, 2 | No change |
| **Language** |  |  |
| Animal fluency | 1, 2 | No change |
| Token test | 1, 2 | No change |
| What is a brown nut that contains milk? | 1 | Retired after wave 1 due to unfamiliarity with coconuts in noncoastal parts of India. |
| What living thing has roots, branches, and leaves? | 2 | Newly added in Wave 2 to replace the coconut item. |
| What is a hammer used for? | 1 |  |
| What is a hammer used for? | 2 | Scoring in wave 2 was expended to allow more flexible responses. |
| What is a sharp object used to cut paper? | 1, 2 | No change |
| Object naming - watch | 1, 2 | No change |
| Object naming - pencil | 1, 2 | No change |
| Elbow naming | 1, 2 | No change |
| Say or write a sentence | 1, 2 | No change |
| Read or copy example "Close your Eyes" | 1, 2 | No change |
| Phrase repetition: Neither this nor that | 1, 2 | No change |
| Point to the floor then ceiling | 1, 2 | No change |
| Local store | 1, 2 | No change |
| **Orientation** |  |  |
| Date naming-date of month | 1, 2 | No change |
| Date naming-month | 1, 2 | No change |
| Date naming-day of week | 1, 2 | No change |
| Date naming-year | 1, 2 | No change |
| Place naming - state | 1, 2 | No change |
| Place naming - city | 1, 2 | No change |
| Date naming-season | 1, 2 | No change |
| What is this place used for? | 1, 2 | No change |
| Place naming-address | 1, 2 | No change |
| Place naming-name of hospital | 1, 2 | Asked in wave 1 when interviews were conducted in facilities |
| Place naming-district of home | 2 | No change |
| Name the prime minister | 1, 2 | No change |
| **Visuospatial** |  |  |
| Constructional praxis, immediate | 1, 2 | No change |
| Figure draw - interlocking pentagons | 1, 2 | No change |
| Clock drawing | 1, 2 | No change |

Legend. LASI-DAD: Longitudinal Aging Study in India – Diagnostic Assessment of Dementia; CERAD: Consortium to Establish a Registry for Alzheimer's Disease

eTable 3. Global fit statistics for general and domain-specific cognitive performance from two-factor Confirmatory Factor Analysis models: Results from LASI-DAD (N=6,168)

| Cognitive domain | RMSEA | CFI | SRMR | Interpretation of fit |
| --- | --- | --- | --- | --- |
| General Cognitive performance | 0.024 | 0.918 | 0.075 | Adequate |
| Memory | 0.034 | 0.948 | 0.045 | Good |
| Executive function/attention | 0.040 | 0.930 | 0.050 | Adequate |
| Language/fluency | 0.025 | 0.902 | 0.075 | Adequate |
| Orientation | 0.029 | 0.975 | 0.073 | Good |
| Visuospatial | 0.070 | 0.969 | 0.037 | Poor |

Legend. Bifactor structures, which corresponded with the same structures used previously in wave 1, were necessary to improve fit to acceptable levels for all domains except visuospatial functioning which had only three indicators. RMSEA: root mean squared error of approximation; CFI: comparative fit index; SRMR: standardized root mean squared residual.

eTable 4. Factor loadings from general and domain-specific confirmatory factor analyses of cognitive performance: Results from LASI-DAD (N=6,168)

| Cognitive test item | Wave in which the variable was measured | Standardized loading, domain-specific factor | Standardized loading, general cognition factor |
| --- | --- | --- | --- |
| **Memory** |  |  |  |
| 3-word registration | 1, 2 | 0.46 | 0.53 |
| 3-word delayed recall | 1, 2 | 0.56 | 0.48 |
| CERAD immediate recall, sum of 3 trials | 1, 2 | 0.8 | 0.67 |
| CERAD delayed word recall | 1, 2 | 0.76 | 0.6 |
| CERAD recognition | 1, 2 | 0.68 | 0.63 |
| Logical Memory immediate, exact | 1, 2 | 0.56 | 0.55 |
| Logical Memory delayed, exact | 1, 2 | 0.55 | 0.55 |
| Logical Memory, recognition | 1, 2 | 0.58 | 0.5 |
| Brave man immediate - Exact | 1, 2 | 0.67 | 0.61 |
| Brave man delayed - Exact | 1, 2 | 0.61 | 0.59 |
| Constructional praxis, delayed | 1, 2 | 0.58 | 0.64 |
| **Executive function/attention** |  |  |  |
| Trails A | 2 | 0.55 | 0.56 |
| Trails B | 2 | 0.3 | 0.32 |
| GoNoGo Part 1 | 1, 2 | 0.7 | 0.68 |
| GoNoGo Part 2 | 1, 2 | 0.66 | 0.64 |
| Similarities and differences | 1, 2 | 0.57 | 0.58 |
| Similarities and differences | 2 | 0.53 | 0.52 |
| DSF | 1, 2 | 0.69 | 0.68 |
| DSB | 1, 2 | 0.83 | 0.81 |
| Symbol Cancellation | 1 | 0.71 | 0.74 |
| Symbol Cancellation | 2 | 0.66 | 0.66 |
| Three-stage task | 1, 2 | 0.49 | 0.5 |
| Serial 7s | 1, 2 | 0.76 | 0.74 |
| Backwards day naming | 1, 2 | 0.74 | 0.75 |
| What to do when raining while out | 2 | 0.55 | 0.56 |
| Computation 1 (sari) | 2 | 0.62 | 0.6 |
| Computation 2 (lottery) | 2 | 0.71 | 0.68 |
| Mental calculation /problem-solving | 1, 2 | 0.74 | 0.73 |
| Ravens | 1, 2 | 0.58 | 0.59 |
| **Language/fluency** |  |  |  |
| Animal fluency | 1, 2 | 0.55 | 0.57 |
| Token test | 1, 2 | 0.57 | 0.59 |
| Coconut | 1 | 0.48 | 0.43 |
| Tree | 2 | 0.63 | 0.55 |
| Hammer | 1 | 0.54 | 0.39 |
| Hammer | 2 | 0.66 | 0.49 |
| Scissors | 1, 2 | 0.61 | 0.46 |
| Object naming - watch | 1, 2 | 0.59 | 0.42 |
| Object naming - pencil | 1, 2 | 0.55 | 0.47 |
| Elbow naming | 1, 2 | 0.6 | 0.43 |
| Say or write a sentence | 1, 2 | 0.57 | 0.48 |
| Read or copy example "Close your Eyes" | 1, 2 | 0.07 | -0.21 |
| Phrase repetition: Neither this nor that | 1, 2 | 0.53 | 0.53 |
| Point to the floor then ceiling | 1, 2 | 0.55 | 0.43 |
| Local store | 1, 2 | 0.72 | 0.59 |
| **Orientation** |  |  |  |
| Date naming-day of week | 1, 2 | 0.52 | 0.59 |
| Date naming-date of month | 1, 2 | 0.67 | 0.65 |
| Date naming-month | 1, 2 | 0.66 | 0.68 |
| Date naming-year | 1, 2 | 0.88 | 0.91 |
| Date naming-season | 1, 2 | 0.52 | 0.48 |
| Place naming - state | 1, 2 | 0.77 | 0.75 |
| Place naming - city | 1, 2 | 0.64 | 0.56 |
| What is this place used for? | 1, 2 | 0.53 | 0.51 |
| Place naming-address | 1, 2 | 0.7 | 0.67 |
| place naming-name of hospital | 1 | 0.75 | 0.65 |
| Place naming-district of home | 2 | 0.69 | 0.59 |
| Name the prime minister | 1, 2 | 0.81 | 0.76 |
| **Visuospatial** |  |  |  |
| Constructional praxis, immediate | 1, 2 | 0.83 | 0.75 |
| Figure draw - interlocking pentagons | 1, 2 | 0.91 | 0.82 |
| Clock drawing | 1, 2 | 0.77 | 0.74 |

eTable 5. Associations of risk factors for dementia with change in memory performance: Results from LASI-DAD (N=2,566)

| Variable | Memory | | | |
| --- | --- | --- | --- | --- |
|  | Beta | SE | z-statistic | Age-stdized beta |
| Demographics and family background |  |  |  |  |
| Age, per 10 years | -0.02 | 0.01 | -3.68 | 10.00 |
| Female sex | -0.01 | 0.01 | -1.86 | 7.22 |
| Any formal schooling | -0.05 | 0.01 | -8.49 | 30.00 |
| Married | 0.01 | 0.01 | 1.71 | -6.11 |
| Scheduled caste or tribe | 0.00 | 0.01 | 0.02 | 0.00 |
| Height, meters | 0.08 | 0.05 | 1.76 | -39.52 |
| Weight, kilograms | 0.00 | 0.00 | -1.40 | 0.00 |
| Body mass index | 0.00 | 0.00 | -2.34 | 0.87 |
| Number of living children | 0.01 | 0.00 | 2.83 | -2.27 |
| Mother's education, any | -0.02 | 0.02 | -0.91 | 7.50 |
| Father's education, any | -0.01 | 0.01 | -1.16 | 6.50 |
| Currently working | 0.01 | 0.01 | 1.88 | -6.67 |
| Health history |  |  |  |  |
| High blood pressure | -0.01 | 0.01 | -2.19 | 7.37 |
| Diabetes | -0.03 | 0.01 | -3.37 | 13.50 |
| Any heart problem | -0.03 | 0.01 | -2.58 | 15.26 |
| Stroke | -0.02 | 0.02 | -1.32 | 12.00 |
| High cholesterol | -0.05 | 0.01 | -3.93 | 23.68 |
| Heart attack | -0.03 | 0.02 | -1.94 | 15.00 |
| Injurious fall | -0.02 | 0.01 | -1.30 | 9.47 |
| Self-reported health and function |  |  |  |  |
| ADL difficulty | -0.02 | 0.01 | -2.25 | 9.44 |
| IADL difficulty | 0.00 | 0.00 | -0.45 | 0.00 |
| Sleep problems | 0.01 | 0.01 | 1.00 | -3.00 |
| Self-rated health (Excellent or good) | 0.01 | 0.01 | 2.20 | -6.19 |
| Probable major depression (CIDI) | 0.01 | 0.01 | 0.63 | -3.64 |
| Objective physical activity |  |  |  |  |
| Balance test | 0.00 | 0.01 | 0.35 | -1.43 |
| Average walking speed (seconds) | 0.00 | 0.00 | -0.34 | 0.45 |
| Health behaviors |  |  |  |  |
| Vigorous activity | 0.01 | 0.01 | 1.47 | -5.26 |
| Moderate activity | -0.01 | 0.01 | -0.92 | 3.00 |
| Alcohol use, past 3 months | 0.00 | 0.01 | 0.06 | -0.50 |
| Currently smokes | 0.03 | 0.01 | 3.13 | -14.21 |
| Sensory function |  |  |  |  |
| Any hearing/ear-related condition | 0.00 | 0.01 | 0.05 | -0.50 |
| Near vision impairment in better eye | 0.01 | 0.01 | 1.12 | -3.04 |
| Distance vision impairment in better eye | 0.00 | 0.01 | 0.25 | -0.87 |

Legend. Shown are regressions of cognitive change on risk factors. All models are adjusted for age, sex, and education. The "Beta" column shows the association of the exposure with change in cognition. The "z-statistic" column is the beta/SE. The column "age-standardized beta" shows the coefficient divided by the age coefficient in that model. SE: standard error; ADL: activities of daily living; IADL: instrumental activities of daily living; CIDI: Composite International Diagnostic Interview.

eTable 6. Associations of risk factors for dementia with change in executive functioning/attention: Results from LASI-DAD (N=2,566)

| Variable | Executive functioning/ attention | | | |
| --- | --- | --- | --- | --- |
|  | Beta | SE | z-statistic | Age-stdized beta |
| Demographics and family background |  |  |  |  |
| Age, per 10 years | -0.01 | 0.00 | -2.97 | 10.00 |
| Female sex | -0.02 | 0.01 | -3.62 | 16.15 |
| Any formal schooling | -0.04 | 0.01 | -7.41 | 30.77 |
| Married | 0.02 | 0.01 | 3.25 | -14.62 |
| Scheduled caste or tribe | 0.01 | 0.01 | 0.74 | -3.13 |
| Height, meters | 0.01 | 0.04 | 0.32 | -6.67 |
| Weight, kilograms | 0.00 | 0.00 | -1.87 | 0.00 |
| Body mass index | 0.00 | 0.00 | -2.32 | 0.53 |
| Number of living children | 0.00 | 0.00 | 1.88 | -1.76 |
| Mother's education, any | -0.03 | 0.02 | -1.82 | 20.00 |
| Father's education, any | -0.03 | 0.01 | -3.38 | 18.33 |
| Currently working | 0.01 | 0.01 | 0.95 | -3.33 |
| Health history |  |  |  |  |
| High blood pressure | -0.01 | 0.01 | -1.69 | 5.63 |
| Diabetes | -0.02 | 0.01 | -2.36 | 10.00 |
| Any heart problem | -0.01 | 0.01 | -1.26 | 6.88 |
| Stroke | -0.01 | 0.02 | -0.86 | 8.75 |
| High cholesterol | -0.01 | 0.01 | -1.00 | 5.63 |
| Heart attack | -0.02 | 0.01 | -1.84 | 13.75 |
| Injurious fall | -0.01 | 0.01 | -0.42 | 3.13 |
| Self-reported health and function |  |  |  |  |
| ADL difficulty | 0.00 | 0.01 | 0.14 | -0.63 |
| IADL difficulty | 0.00 | 0.00 | 0.85 | -0.56 |
| Sleep problems | 0.01 | 0.01 | 2.34 | -7.06 |
| Self-rated health (Excellent or good) | 0.00 | 0.01 | 0.78 | -2.35 |
| Probable major depression (CIDI) | 0.02 | 0.01 | 1.85 | -10.59 |
| Objective physical activity |  |  |  |  |
| Balance test | 0.01 | 0.01 | 1.14 | -4.12 |
| Average walking speed (seconds) | 0.00 | 0.00 | -0.01 | 0.00 |
| Health behaviors |  |  |  |  |
| Vigorous activity | 0.00 | 0.01 | 0.11 | -0.63 |
| Moderate activity | 0.00 | 0.01 | -0.01 | 0.00 |
| Alcohol use, past 3 months | 0.02 | 0.01 | 1.70 | -11.25 |
| Currently smokes | 0.01 | 0.01 | 1.68 | -8.13 |
| Sensory function |  |  |  |  |
| Any hearing/ear-related condition | 0.03 | 0.01 | 2.72 | -13.89 |
| Near vision impairment in better eye | 0.00 | 0.01 | -0.55 | 1.67 |
| Distance vision impairment in better eye | 0.01 | 0.01 | 1.70 | -5.00 |

Legend. Shown are regressions of cognitive change on risk factors. All models are adjusted for age, sex, and education. The "Beta" column shows the association of the exposure with change in cognition. The "z-statistic" column is the beta/SE. The column "age-standardized beta" shows the coefficient divided by the age coefficient in that model. SE: standard error; ADL: activities of daily living; IADL: instrumental activities of daily living; CIDI: Composite International Diagnostic Interview.

eTable 7. Associations of risk factors for dementia with change in language: Results from LASI-DAD (N=2,566)

| Variable | Language/fluency | | | |
| --- | --- | --- | --- | --- |
|  | Beta | SE | z-statistic | Age-standardized beta |
| Demographics and family background |  |  |  |  |
| Age, per 10 years | -0.01 | 0.01 | -1.91 | 10.00 |
| Female sex | -0.01 | 0.01 | -0.64 | 4.55 |
| Any formal schooling | -0.05 | 0.01 | -6.68 | 41.82 |
| Married | 0.02 | 0.01 | 3.00 | -20.91 |
| Scheduled caste or tribe | -0.01 | 0.01 | -0.87 | 4.67 |
| Height, meters | -0.06 | 0.05 | -1.23 | 30.00 |
| Weight, kilograms | 0.00 | 0.00 | -1.88 | 0.48 |
| Body mass index | 0.00 | 0.00 | -1.61 | 0.50 |
| Number of living children | 0.01 | 0.00 | 3.68 | -3.89 |
| Mother's education, any | -0.03 | 0.02 | -1.47 | 21.33 |
| Father's education, any | -0.01 | 0.01 | -1.07 | 8.00 |
| Currently working | 0.00 | 0.01 | 0.41 | -2.00 |
| Health history |  |  |  |  |
| High blood pressure | -0.02 | 0.01 | -2.82 | 13.57 |
| Diabetes | -0.02 | 0.01 | -1.87 | 10.67 |
| Any heart problem | 0.00 | 0.01 | -0.02 | 0.00 |
| Stroke | -0.01 | 0.03 | -0.46 | 8.00 |
| High cholesterol | -0.02 | 0.01 | -1.75 | 15.33 |
| Heart attack | 0.00 | 0.02 | -0.07 | 0.67 |
| Injurious fall | 0.00 | 0.02 | -0.19 | 5.00 |
| Self-reported health and function |  |  |  |  |
| ADL difficulty | 0.00 | 0.01 | -0.06 | 0.67 |
| IADL difficulty | 0.00 | 0.00 | 0.51 | 0.00 |
| Sleep problems | 0.01 | 0.01 | 1.95 | -8.67 |
| Self-rated health (Excellent or good) | 0.01 | 0.01 | 1.25 | -4.71 |
| Probable major depression (CIDI) | 0.03 | 0.01 | 2.28 | -17.78 |
| Objective physical activity |  |  |  |  |
| Balance test | 0.01 | 0.01 | 1.24 | -5.56 |
| Average walking speed (seconds) | 0.00 | 0.00 | -0.36 | 0.40 |
| Health behaviors |  |  |  |  |
| Vigorous activity | 0.01 | 0.01 | 1.45 | -7.86 |
| Moderate activity | 0.00 | 0.01 | -0.37 | 2.00 |
| Alcohol use, past 3 months | 0.01 | 0.01 | 0.44 | -3.33 |
| Currently smokes | 0.01 | 0.01 | 0.92 | -6.00 |
| Sensory function |  |  |  |  |
| Any hearing/ear-related condition | 0.02 | 0.01 | 1.65 | -12.50 |
| Near vision impairment in better eye | 0.01 | 0.01 | 1.31 | -4.50 |
| Distance vision impairment in better eye | 0.01 | 0.01 | 0.86 | -3.33 |

Legend. Shown are regressions of cognitive change on risk factors. All models are adjusted for age, sex, and education. The "Beta" column shows the association of the exposure with change in cognition. The "z-statistic" column is the beta/SE. The column "age-standardized beta" shows the coefficient divided by the age coefficient in that model. SE: standard error; ADL: activities of daily living; IADL: instrumental activities of daily living; CIDI: Composite International Diagnostic Interview.

eTable 8. Associations of risk factors for dementia with change in orientation: Results from LASI-DAD (N=2,566)

| Variable | Orientation | | | |
| --- | --- | --- | --- | --- |
|  | Beta | SE | z-statistic | Age-standardized beta |
| Demographics and family background |  |  |  |  |
| Age, per 10 years | -0.01 | 0.00 | -2.58 | 10.00 |
| Female sex | -0.01 | 0.01 | -1.66 | 8.33 |
| Any formal schooling | -0.03 | 0.01 | -5.43 | 25.00 |
| Married | 0.01 | 0.01 | 1.03 | -5.00 |
| Scheduled caste or tribe | 0.00 | 0.01 | -0.11 | 0.77 |
| Height, meters | -0.07 | 0.04 | -1.85 | 55.38 |
| Weight, kilograms | 0.00 | 0.00 | -2.10 | 0.00 |
| Body mass index | 0.00 | 0.00 | -1.59 | 0.83 |
| Number of living children | 0.00 | 0.00 | 2.57 | -2.67 |
| Mother's education, any | 0.01 | 0.02 | 0.29 | -3.85 |
| Father's education, any | -0.01 | 0.01 | -0.93 | 6.92 |
| Currently working | 0.00 | 0.01 | 0.06 | 0.00 |
| Health history |  |  |  |  |
| High blood pressure | -0.01 | 0.01 | -2.14 | 9.17 |
| Diabetes | -0.02 | 0.01 | -2.15 | 11.54 |
| Any heart problem | -0.01 | 0.01 | -1.52 | 10.77 |
| Stroke | -0.03 | 0.02 | -1.51 | 20.00 |
| High cholesterol | -0.02 | 0.01 | -1.70 | 13.08 |
| Heart attack | -0.03 | 0.01 | -2.29 | 23.85 |
| Injurious fall | -0.01 | 0.01 | -1.08 | 26.00 |
| Self-reported health and function |  |  |  |  |
| ADL difficulty | 0.00 | 0.01 | -0.48 | 2.50 |
| IADL difficulty | 0.00 | 0.00 | -0.06 | 0.00 |
| Sleep problems | 0.01 | 0.01 | 1.32 | -5.38 |
| Self-rated health (Excellent or good) | 0.00 | 0.01 | 0.35 | -1.54 |
| Probable major depression (CIDI) | 0.02 | 0.01 | 2.36 | -16.43 |
| Objective physical activity |  |  |  |  |
| Balance test | 0.01 | 0.01 | 1.30 | -9.00 |
| Average walking speed (seconds) | 0.00 | 0.00 | 0.22 | 0.00 |
| Health behaviors |  |  |  |  |
| Vigorous activity | 0.01 | 0.01 | 0.82 | -3.85 |
| Moderate activity | -0.01 | 0.01 | -1.74 | 6.43 |
| Alcohol use, past 3 months | 0.03 | 0.01 | 2.71 | -22.50 |
| Currently smokes | 0.02 | 0.01 | 2.12 | -11.54 |
| Sensory function |  |  |  |  |
| Any hearing/ear-related condition | 0.02 | 0.01 | 1.58 | -10.71 |
| Near vision impairment in better eye | 0.01 | 0.01 | 1.27 | -5.38 |
| Distance vision impairment in better eye | 0.00 | 0.01 | 0.45 | -2.31 |

Legend. Shown are regressions of cognitive change on risk factors. All models are adjusted for age, sex, and education. The "Beta" column shows the association of the exposure with change in cognition. The "z-statistic" column is the beta/SE. The column "age-standardized beta" shows the coefficient divided by the age coefficient in that model. SE: standard error; ADL: activities of daily living; IADL: instrumental activities of daily living; CIDI: Composite International Diagnostic Interview.

eTable 9. Associations of risk factors for dementia with change in visuospatial function: Results from LASI-DAD (N=2,566)

| Variable | Visuospatial function | | | |
| --- | --- | --- | --- | --- |
|  | Beta | SE | z-statistic | Age-stdized beta |
| Demographics and family background |  |  |  |  |
| Age, per 10 years | -0.01 | 0.00 | -2.35 | 10.00 |
| Female sex | 0.02 | 0.01 | 2.93 | -18.00 |
| Any formal schooling | 0.00 | 0.01 | -0.48 | 3.00 |
| Married | -0.01 | 0.01 | -1.64 | 10.00 |
| Scheduled caste or tribe | -0.01 | 0.01 | -1.61 | 12.50 |
| Height, meters | -0.04 | 0.04 | -0.83 | 32.73 |
| Weight, kilograms | 0.00 | 0.00 | -0.82 | 0.00 |
| Body mass index | 0.00 | 0.00 | -0.53 | 0.00 |
| Number of living children | 0.00 | 0.00 | -0.18 | 0.00 |
| Mother's education, any | 0.00 | 0.02 | -0.09 | 2.22 |
| Father's education, any | -0.01 | 0.01 | -1.18 | 11.00 |
| Currently working | -0.01 | 0.01 | -1.83 | 11.00 |
| Health history |  |  |  |  |
| High blood pressure | -0.01 | 0.01 | -0.91 | 6.25 |
| Diabetes | 0.00 | 0.01 | 0.43 | -3.75 |
| Any heart problem | 0.01 | 0.01 | 0.68 | -8.75 |
| Stroke | 0.04 | 0.02 | 2.02 | -52.50 |
| High cholesterol | 0.01 | 0.01 | 1.05 | -15.00 |
| Heart attack | 0.02 | 0.01 | 1.28 | -22.50 |
| Injurious fall | -0.01 | 0.01 | -0.43 | 10.00 |
| Self-reported health and function |  |  |  |  |
| ADL difficulty | -0.01 | 0.01 | -0.83 | 7.50 |
| IADL difficulty | 0.00 | 0.00 | 1.89 | -0.91 |
| Sleep problems | 0.01 | 0.01 | 1.68 | -11.25 |
| Self-rated health (Excellent or good) | -0.01 | 0.01 | -0.99 | 5.56 |
| Probable major depression (CIDI) | -0.01 | 0.01 | -1.18 | 13.33 |
| Objective physical activity |  |  |  |  |
| Balance test | 0.01 | 0.01 | 1.27 | -9.00 |
| Average walking speed (seconds) | 0.00 | 0.00 | 1.87 | -2.00 |
| Health behaviors |  |  |  |  |
| Vigorous activity | -0.02 | 0.01 | -2.56 | 17.00 |
| Moderate activity | -0.01 | 0.01 | -1.47 | 8.89 |
| Alcohol use, past 3 months | 0.00 | 0.01 | 0.20 | -2.50 |
| Currently smokes | 0.02 | 0.01 | 2.43 | -28.57 |
| Sensory function |  |  |  |  |
| Any hearing/ear-related condition | 0.01 | 0.01 | 1.08 | -11.11 |
| Near vision impairment in better eye | 0.01 | 0.01 | 1.59 | -8.33 |
| Distance vision impairment in better eye | 0.01 | 0.01 | 0.77 | -4.17 |

Legend. Shown are regressions of cognitive change on risk factors. All models are adjusted for age, sex, and education. The "Beta" column shows the association of the exposure with change in cognition. The "z-statistic" column is the beta/SE. The column "age-standardized beta" shows the coefficient divided by the age coefficient in that model. SE: standard error; ADL: activities of daily living; IADL: instrumental activities of daily living; CIDI: Composite International Diagnostic Interview.

eFigure 1. Histograms of general and domain-specific cognitive performance by wave: Results from LASI-DAD (N=6,168)

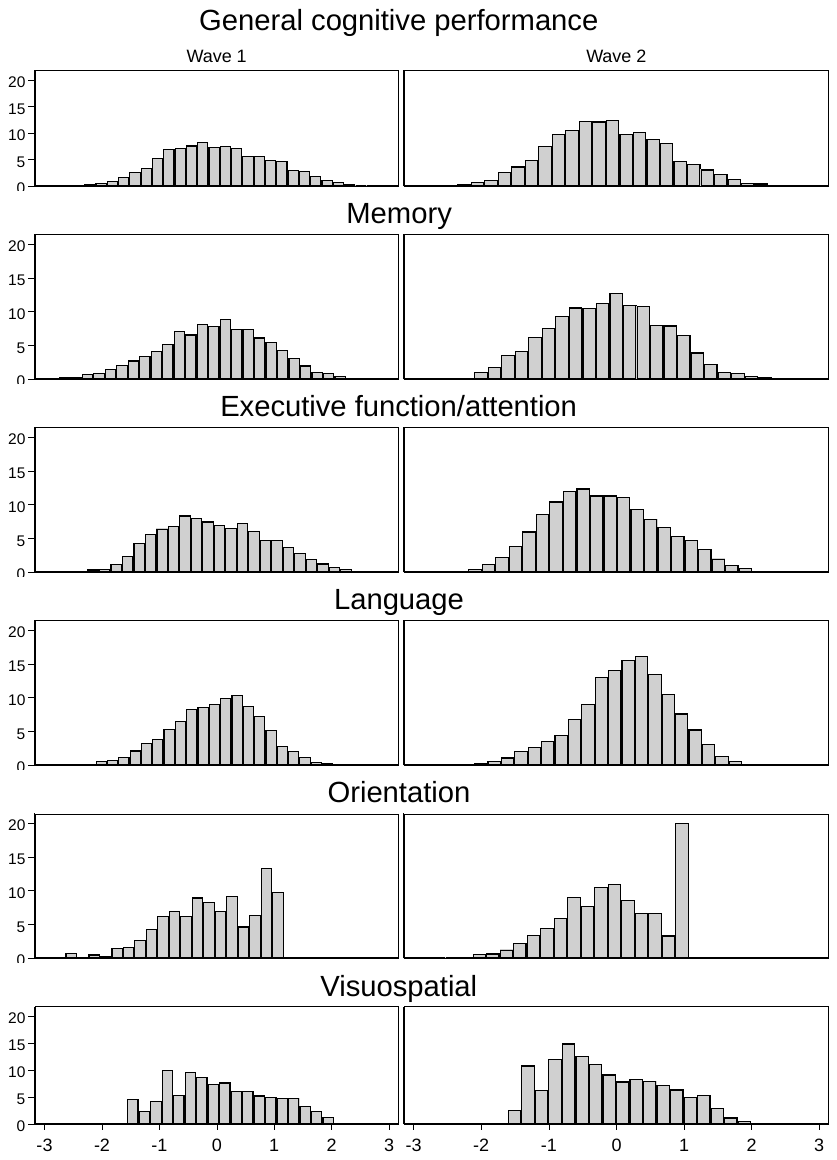

Legend. Bar heights in each histogram indicate relative frequency of cognitive domain scores with a value along the x-axis.

eFigure 2. Marginal reliability of general and domain-specific cognitive performance: Results from LASI-DAD (N=6,168)

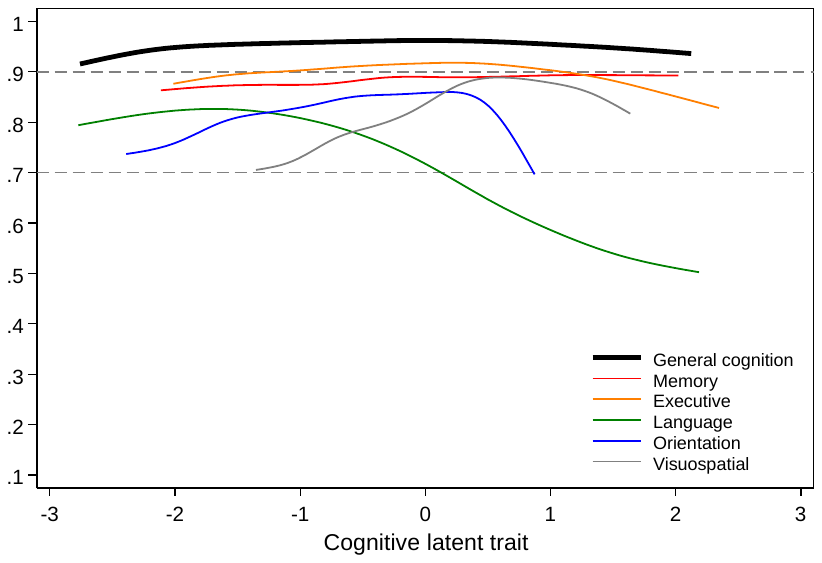

Legend. This figure shows differences in the reliability of the estimation model for each cognitive domain as a function of corresponding abilities on the latent trait. Horizontal dashed lines at reliabilities of 0.7 and 0.9 illustrate acceptable thresholds of reliability for basic research and high-stakes testing, respectively.
